# Melanoma Skin Cancer Detection using Deep Learning

**DOI:** 10.1101/2022.02.02.22269505

**Authors:** Dheiver Santos

**Affiliations:** BRIDGE – Instituto de Tecnologia e Pesquisa, Estado / UF: Sergipe / SE Município: Aracaju Bairro: Getúlio Vargas Logradouro: Rua São Cristóvão Número: 1361 Complemento: CEP: 49.055-620; Faculdade Estácio - Estácio de Sá Av. Pio XII, 350 - Jatiúca, Maceió - AL, 57035-560

## Abstract

Data from the World Health Organization (WHO) indicate a worldwide occurrence of 2 to 3 million cases of non-melanoma skin cancer annually. The American Cancer Society estimates that the incidence reaches 5.4 million in the United States alone. In cases of fatal diseases, early detection received great attention from the population and the media due to the premise that the earlier a cancer is identified, the greater the chances of cure. It is to be believed that the application of automated methods will help in early diagnosis, especially with the set of images with a variety of diagnoses. Thus, this article presents a system for recognizing dermatological diseases through images with lesions, a machine intervention in contrast to conventional detection based on medical personnel. Our model is designed in three phases, committing to data collection and augmentation, model development, and finally, prediction. We used various AI algorithms such as ANN with image processing tools to form a better structure, leading to higher accuracy of 89%.

## 1.0 Introduction

According to the most recent data from the World Health Organization (WHO), cancer is the second leading cause of death in the world and was responsible for 9.6 million deaths in 2018. Globally, one in six deaths is related to the disease and approximately 70% of cancer deaths occur in low- and middle-income countries.[1,2] The presentation of late and inaccessible diagnosis and treatment are common. In 2017, only 26% of low-income countries reported having pathology services available in the public sector; on the other hand, more than 90% of high-income countries reported that treatment services are available, compared to less than 30% in low-income countries. [1,3] The economic impact of cancer is significant and increasing. In the last global survey, the total annual cost of the disease in 2010 was estimated at approximately US$ 1.16 trillion (PAHO, 2018).

Imaging tests help to locate the lesion and are extremely useful in determining the extent of the disease, which is called the staging of cancer. Due to the large volume of information generated by this type of image, these technologies have drawn attention in research areas related to Computer Science, with studies on the analysis of these images with the aid of Artificial Intelligence techniques. [4,5,6]

Thus, among the most implemented techniques in Intelligence Artificial, the ANN with Deep Learning architecture stands out, in which, due to their robustness and the quality of the results obtained, have been widely used in applications that handle large volumes of data (Big Data, for example), and in special for Digital Image Processing. In this way, the ANN with the architecture of Deep Learning presented itself as a promising option for the application in this work.

## 2.0 Dataset

The dataset used in this study consists of 2357 images of various oncologic malignancies and malignancies developed by the International Skin Imaging Association (ISIC)[7]. All images were classified according to the classification constructed by the ISIC, and all subcategories were divided into some images, except for melanomas and moles in which images predominated. In Figures 1. and 2, the reader is presented with the diseases that will be identified by the neural network.

**Figure 1.**
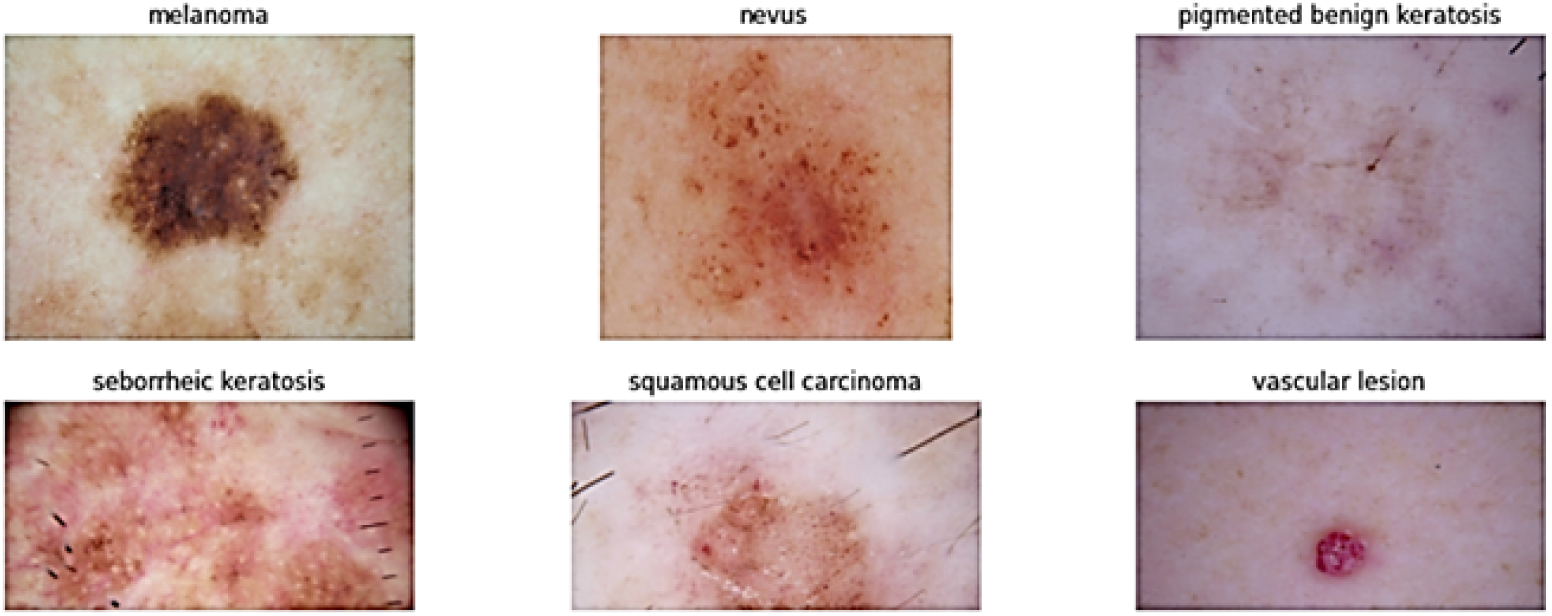
Skin Cancer types Example

**Figure 2.**
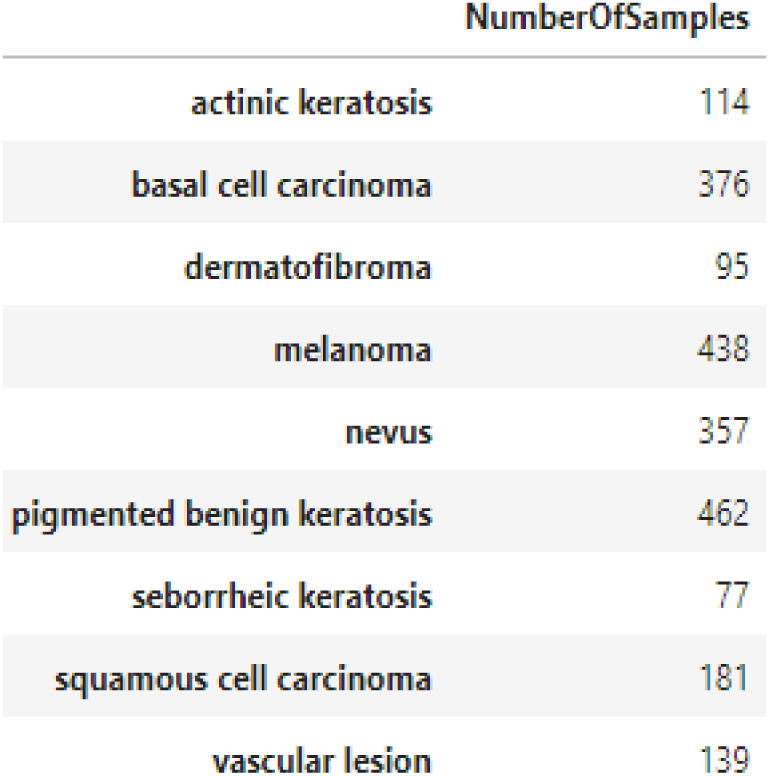
Skin Cancer Samples

## 3.0 Deep Learning

Artificial Neural Networks are mathematical computational techniques capable of solving problems through simple circuits that simulate the functioning and behavior of the human brain [4,5]. One of the great applications of artificial neural networks is its use for pattern recognition, having been used for the identification of cancer cells. A neural network is a parallel and distributed system, formed by simple processing units, which has the ability to store knowledge from the learning process in which connection forces between neurons known as synaptic weights are used to store the acquired knowledge and make it available for use 5,6].

You can see the architecture and layers of the discriminator and generator network in image 3. In the Conv2D layers of the discriminator and in the Conv2DTranspose and Conv2D layers of the generator, padding: same was used. Next to the params values, when indicated, the latent space (EL) values appear in parentheses.

The Conv2DTranspose (Transposed Convolution Layer) layers work like an inverse convolution, expanding the dimensions of the data. In older literature it is called the Deconvolution Layer.

### CNN Architecture

The proposed CNN architecture. The input layer of the neural network is becoming a 64 × 64 pixel RGB image. The input data group (images) are processed by three convolutional blocks. Each convolutional block contains 2D convolution layer operation. Any hidden layer includes a ReLU (Rectified Linear Unit) as the activation function layer (nonlinear operation) and is prepared with spatial pooling using a max-pooling layer.

The network is complete with a classifier block, composed of two fully-connected layers. Softmax function is required as a classifier for a fully linked layer output. Our network makes it possible to decide whether the lesion is malignant and benign tumors. The architecture for the network model is defined in Figure 3.

**Figure 3.**
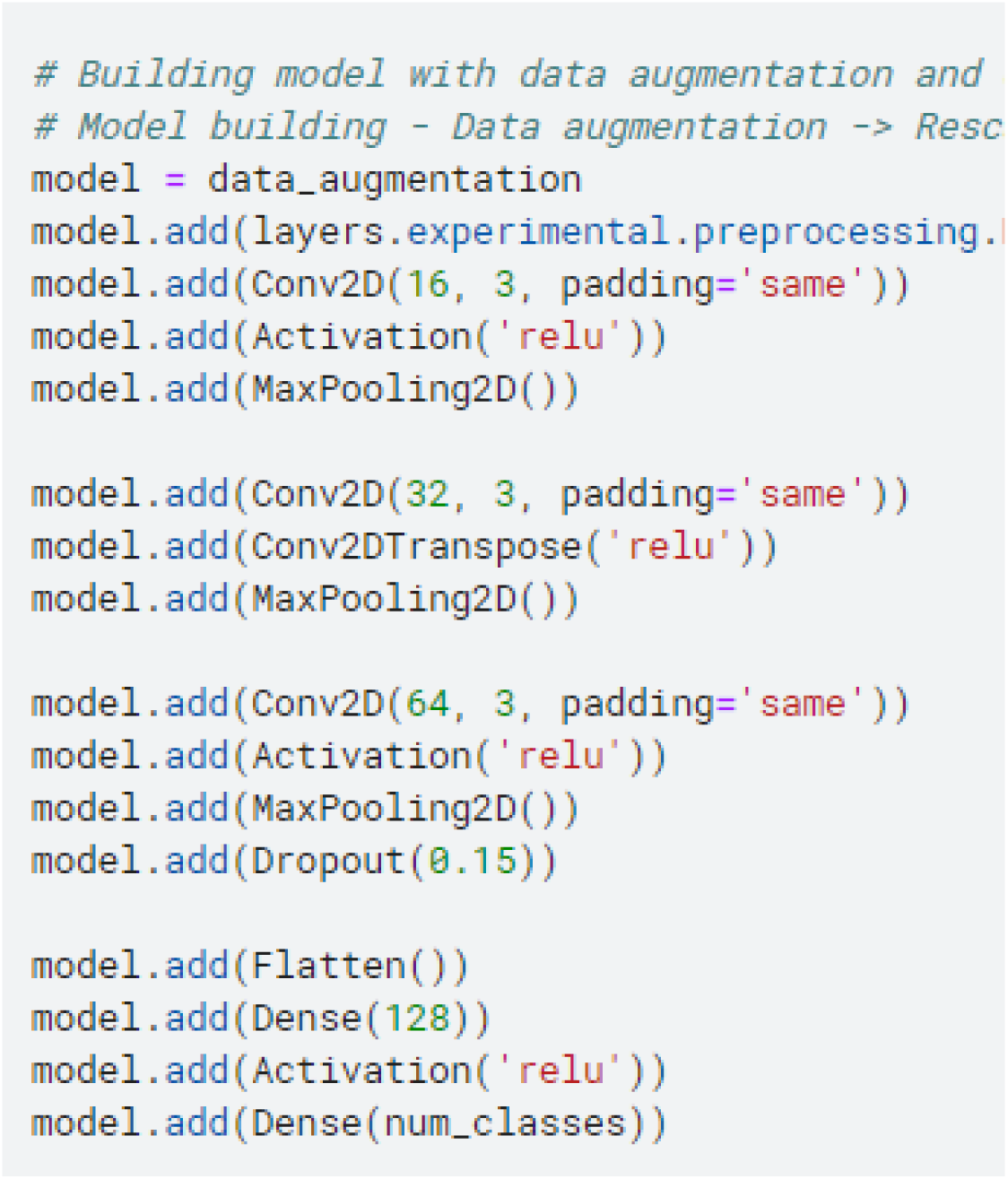
CNN Architecture

### Training

The proposed neural network is trained based on the “Adam” optimization using the learning rate of 2e-4. In this work, the number of epochs is 100, a value of 0.5 is used for a dropout optimization on fully connected layers and the batch size is set to 32.

## 4.0 Results

The images are then added inside the dataframe. The image dataset is divided into training, validation, and testing datasets. Figure 4 represents the accuracy and loss obtained when the ANN model is applied to the training and test dataset when the ANN model is applied to the training data for fifty epochs loss: 0.1294 - accuracy: 0.9532 - val_loss: 0.5125 - val_accuracy: 0.8889.

**Figure 4.**
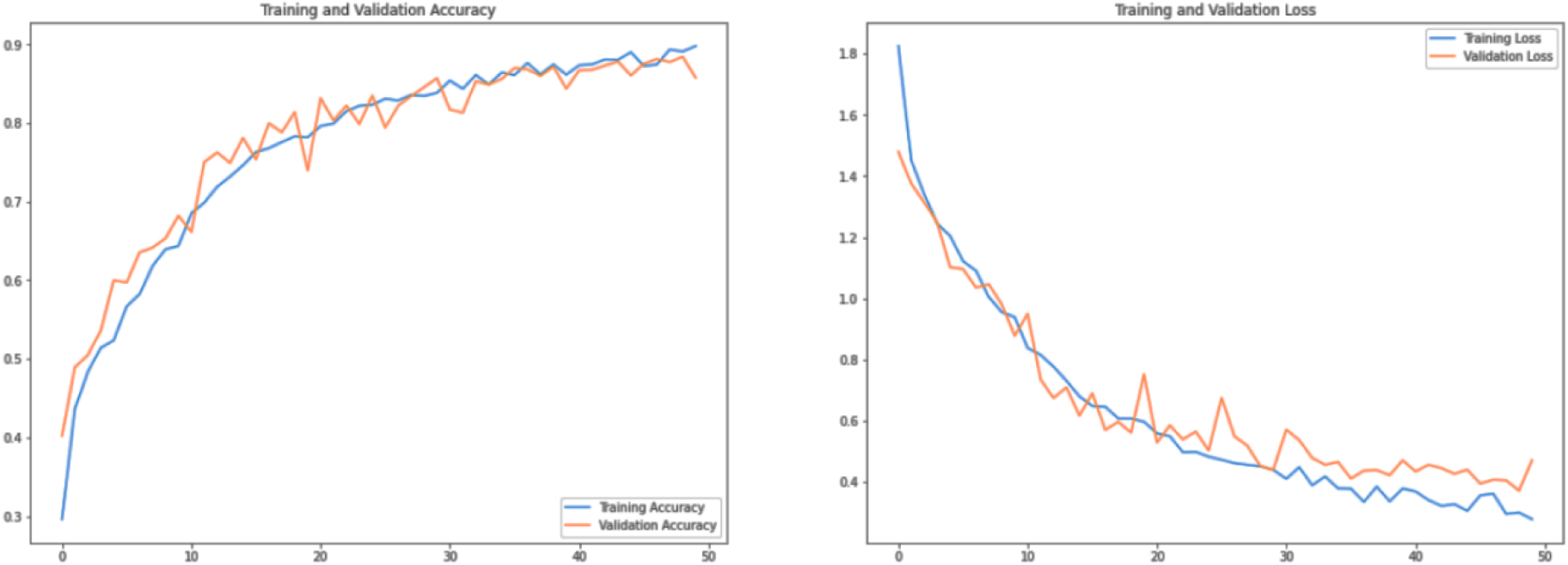
Comparing training/validation accuracy and loss of ANN model.

In Figure 5, it was possible to observe the classification of the typology related to cancer. Reminding readers that all cases are of cancer, what we want is to classify the typology, for example, to know if a certain image types A or B. Our results predict an 89% chance of correctly classifying the type of cancer-based on the said processed dataset.

**Figure 5.**
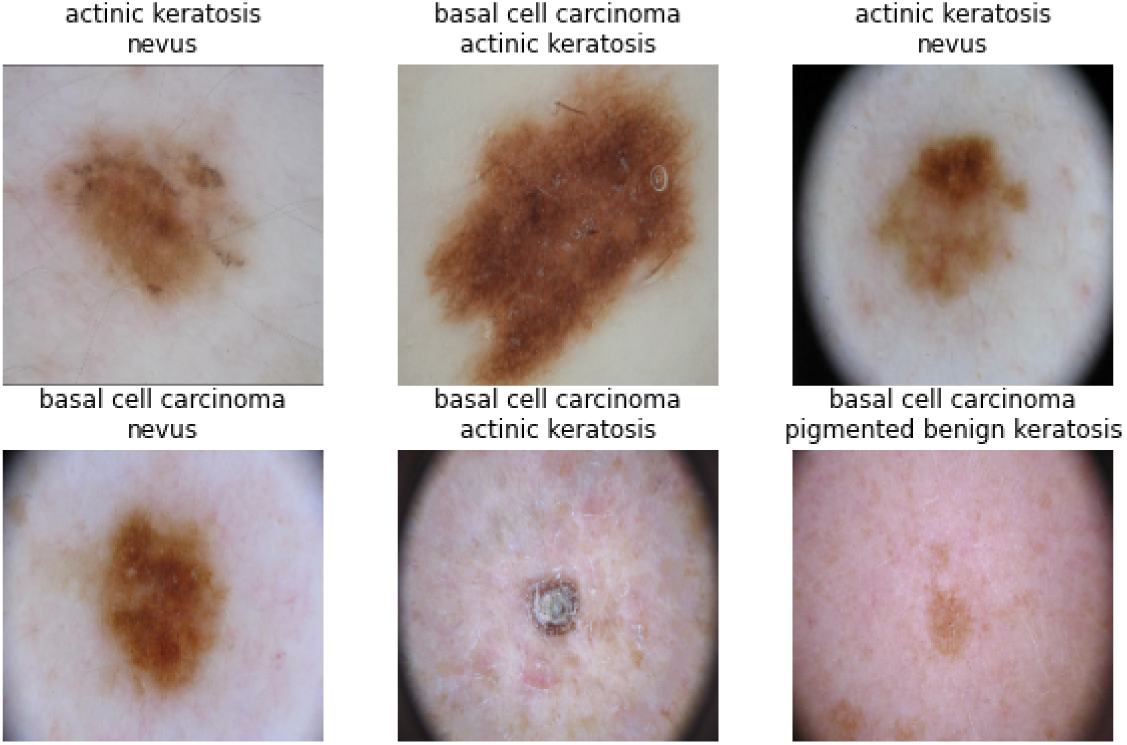
Result test dataset classification

## 5.0 Conclusion

The work proposes to develop a method capable of classifying cancer images from a classic ISIC database. The images are of different types of skin cancer. The work used Deep Learning with different layers to make this classification of a test dataset (data that does not have labels or classification). The results of the metric established an impressive 89% accuracy level. The work can be used by companies for the development of new products or services that help health professionals make faster decisions.

## Data Availability

All data produced in the present study are available upon reasonable request to the authors

